# Premature Atrial Stimulation Accentuates Conduction Abnormalities in Cardiac Surgery Patients that Develop Postoperative Atrial Fibrillation

**DOI:** 10.1101/2021.07.29.21261342

**Authors:** Muhammad S. Khan, Matthias Lange, Ravi Ranjan, Vikas Sharma, Jason P. Glotzbach, Craig Selzman, Derek J. Dosdall

## Abstract

**BACKGROUND:** postoperative atrial fibrillation (POAF) is a common cardiac surgery complication that is associated with increased complications and negative outcomes, but the association between presurgical atrial conduction abnormalities and POAF has not been investigated clinically during premature atrial S1S2 stimulation. This clinical study sought to examine whether intraoperative premature atrial stimulation reveals increased areas of slowed and/or blocked conduction in patients that develop POAF.

**METHODS:** High-density intraoperative epicardial left atrial mapping was conducted in 20 cardiac surgery patients with no prior history of atrial fibrillation (AF). In 20 patients, 6 (30%) developed POAF. A flexible-array of 240-electrodes was placed on the posterior left atrial wall in between the pulmonary veins. Activation maps were generated for sinus and premature atrial S1S2 stimulated beats. The area of conduction block (CB), conduction delay (CD) and the combination of both (CDCB) for conduction velocity <0.1, 0.1≤x<0.2 and <0.2 m/s, respectively were quantified.

**RESULTS:** For a premature atrial S2 beat with shortest cycle length captured, conduction velocity maps revealed a significantly higher area for CD (13.19±6.59 versus 6.06±4.22 mm^2^, p=0.028) and CDCB (17.36±8.75 versus 7.41±6.39 mm^2^, p=0.034), and a trend toward a larger area for CB (4.17±3.66 versus 1.34±2.86 mm^2^, p=0.063) in patients who developed POAF in comparison to those that remained in the sinus. Sinus and S1 paced beats did not show substantial differences in abnormal conduction areas between patients with and without POAF.

**CONCLUSION:** In comparison to sinus and S1 beats, premature atrial S2 beats accentuate conduction abnormalities in cardiac surgery patients that developed POAF.

## INTRODUCTION

Postoperative atrial fibrillation (POAF) has been associated with increased morbidity and mortality after cardiac surgery [1]. In cardiac surgery patients that experienced an arrhythmia, 70% developed POAF before the end of the fourth postoperative day and 94% before the end of the sixth postoperative day [2]. Treatment of POAF is estimated to add an additional $1 billion in health-care costs in the US alone [3]. Several algorithms based on patients’ demographics and clinical co-morbidities have been developed to forecast complications of following cardiac surgery, but these algorithms lack precision to accurately predict an individual patient’s risk [4,5]. Recent literature has revealed many biomarkers that might be associated with POAF risk [6]. However, significant variation has been noted within biomarkers of interest [7,8].

Conduction abnormalities such as conduction block (CB), conduction delay (CD), and the combination of both (CDCB) play a crucial role in both genesis and perpetuation of atrial arrhythmias in cardiac surgery patients with and without history of atrial fibrillation (AF) [9]. Conduction abnormalities during sinus rhythm (SR) mainly occur at specific locations of nonuniform anisotropic tissue properties due to interstitial fibrosis [10,11]. The association between conduction disorders and the POAF incidence during sinus has previously been studied in the area between right and left pulmonary veins (PV) of cardiac surgery patients [12–14]. In one study, there was no correlation of conduction abnormalities with POAF development [11], whereas the same group later reported a trend towards association of CB and CD lines present at the PV area with POAF incidence in a subset of 70 patients [15].

Previously it has been reported that a premature atrial stimulus creates impulses propagation in a substrate with more dispersed refractoriness and provokes arrhythmogenic conduction in human atrial bundles [16]. Further, atrial conduction disorders increase during spontaneous atrial extra-systoles, which are caused by anisotropy due to their ectopic origin and/or refractoriness of myocardium [17]. This shows that activation delay increases with progressively shorter coupling intervals, and the resulting abnormal conduction patterns are associated with depressed action potentials. Therefore, we envision that a premature atrial stimulus may show an association between preexisting atrial conduction abnormalities and post-surgery AF incidence. In this prospective study of 20 patients, the primary objective is to investigate whether conduction abnormalities (CB, CD and CDCB) for a premature atrial stimulus S2 beat is effective in revealing a pre-surgical atrial substrate that may provide a new method with an increased accuracy for evaluating the risk of POAF in cardiac surgery patients. The conduction disorder data quantified for premature atrial S2 beats are also compared with sinus and S1 beats in group of patients who developed POAF and in those who remained in sinus following cardiac surgery.

## MATERIALS AND METHODS

### Study Patients

The study population consisted of total 22 adult patients with no previous history of AF undergoing first time elective coronary artery bypass grafting, aortic or mitral valve surgery, or a combination of these procedures, between April 2018 and January 2020. The study protocol was approved by the Institutional Review Board of the University of Utah – Health (IRB_00101187). All experiments were performed in accordance with relevant guidelines and regulations as approved, and the written informed consent was obtained from all 22 patients. Two patients were later excluded from the study due to failure of equipment to send S2 stimulation to create premature beats. From 20 patients, POAF incidence was reported in 6 (30%) patients within 1 – 4 days of cardiac surgery. Patients demographics, baseline characteristics, clinical data, lab results and echocardiography measures and intraoperative values were extracted from electronic patients records and are summarized in **Table 1**. The summary of the enrolled patients and intraoperative conduction parameters quantified in this prospective study is shown in **Fig. 1**.

**Table 1.** Demographics and clinical characteristics of cardiac surgery patients.

|  | **All Patients**<br>**(N=20)** | **Sinus**<br>**(n=14)** | **POAF**<br>**(n=6)** | **p value** |
| --- | --- | --- | --- | --- |
| Age (years), M±SD | 61±15 | 57±16 | 71±6 | 0.054 |
| Male, n/N (%) | 14/20 (70) | 10/14 (71) | 4/6 (67) | 0.842 |
| **Cardiovascular Risk factors** |  |  |  |  |
| BMI (kg/m<sup>2</sup>), M±SD | 36.4±8.5 | 38.3±9.2 | 32.1±4.9 | 0.140 |
| Diabetes (m<sup>2</sup>), n/N (%) | 8/20 (40) | 4/14 (29) | 4/6 (67) | 0.161 |
| Smoking, n/N (%) | 11/20 (55) | 8/14 (57) | 3/6 (50) | 0.783 |
| Alcohol, n/N (%) | 10/20 (50) | 8/14 (57) | 2/6 (33) | 0.355 |
| Hypertension, n/N (%) | 14/20 (70) | 9/14 (64) | 5/6 (83) | 0.421 |
| Hyperlipidemia, n/N (%) | 13/20 (65) | 8/14 (57) | 5/6 (83) | 0.285 |
| **Echocardiography** |  |  |  |  |
| LA area (cm<sup>2</sup>), M±SD | 20.2±3.4 | 21.1±3.5 | 17.8±1.5 | 0.089 |
| RA area (cm<sup>2</sup>), M±SD | 17.5±5.3 | 18±6 | 16.1±2.8 | 0.545 |
| EF (%), >50%, n/N (%) | 19/20 | 13/14 (93) | 6/6 (100) | -- |
| **Preoperative Medications** |  |  |  |  |
| β-blockers, n/N (%) | 11/20 (55) | 7/14 (50) | 4/6 (67) | 0.518 |
| Ca channel blockers, n/N | 4/20 (20) | 4/14 (29) | 0/6 (0) | 0.265 |
| ACEI, n/N (%) | 6/20 (30) | 3/14 (21) | 2/6 (33) | 0.597 |
| ARB, n/N (%) | 1/20 (0.05) | 1/14 (7) | 0/6 (0) | -- |
| Statins, n/N (%) | 12/20 (60) | 8/14 (57) | 4/6 (67) | 0.709 |
| Aspirin, n/N (%) | 8/20 (40) | 6/14 (43) | 2/6 (33) | 0.708 |
| Diuretics, n/N (%) | 5/20 (25) | 4/14 (29) | 1/6 (17) | 0.597 |
| **Intraoperative Conditions** |  |  |  |  |
| Pump time (min), M±SD | 153.4±61.5 | 153.7±49 | 152.2±90.3 | 0.969 |
| XCL (min), M±SD | 104.8±38 | 110.86±42.7 | 90.5±20.4 | 0.285 |

**Fig. 1.**
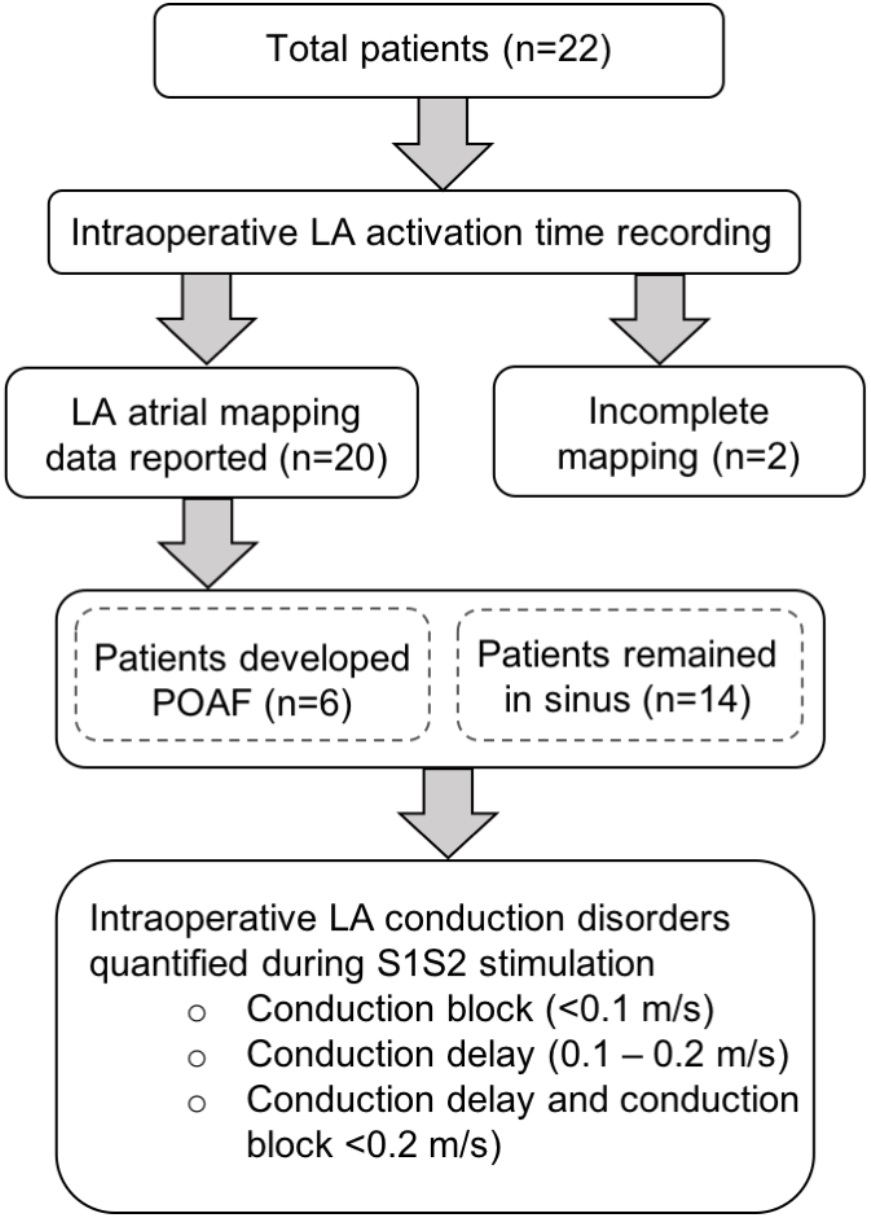
Patients included in the prospective study and quantified conduction parameters.

### Flexible Array of Electrodes and Pacing Protocols

A custom-made, high-density rectangular flexible array of microelectrodes (240 gold plated electrodes with an interelectrode distance of 2.5 mm) was placed on the posterior left atrial (LA) wall in between the PV (**Fig. S1)**. For each patient, the array of electrodes was positioned by cardiothoracic surgeon after bypass cannula placement, and sinus data were evaluated for continuous 30 – 90 sec to ensure the array was in direct contact with atria as opposed to the ventricles. A drive train of 8 S1 beats at a cycle length (CL) of 600 ms was followed by a single premature atrial S2 beat coupled at 400 ms, and reduced in 20 ms steps to 300 ms, then in 10 ms steps until the S2 atrial capture was lost. The biphasic pulse width for this protocol was 2 ms at a current of twice the minimum threshold current of atrial capture. Atrial electrograms were recorded for sinus beats and during premature S1S2 atrial pacing at the same LA location. The pacing protocols were completed before putting the patients into cardiopulmonary bypass.

### Epicardial Activation Time Maps and Postoperative Data Analysis

Isochronal maps were generated for SR beats, for the last S1 beat of consecutive 8 beats with each CL of 600 ms, and also for the shortest pacing CL for the last premature beat S2 at which the atrial beat was successfully captured. Isochronal maps were generated in local activation time (AT) to assess LA activation patterns. Due to time constraints that rendered repositioning of the plaque unfeasible to find the best pacing sites, only data from pacing sites with stable capture were considered for analysis. Recording sites with >25% electrograms missing were not included in the data analysis (n=3). In 3 patients, following S1S2 stimulation, the atrial beat was successfully captured for all S1 at CL 600 ms but not for S2 during CL 400 – 160 ms. All electrograms were recorded and stored on a computerized mapping system (Active Two, BioSEMI B.V., Amsterdam, Netherlands) for offline analysis. Local AT maps were constructed for SR, S1 and S2 beats through the steepest negative slope of atrial potentials recorded for each electrode. We modified the open-source software platform (version 1), ‘RHYTHM’ [18] to analyze recorded AT mapping data. In this toolbox, the data can be filtered, a multipoint derivative can be taken, and AT on each electrode can also be calculated.

### Regional Conduction Velocity and Conduction Disorders

Conduction velocity maps were accessed using the phase mapping method as described elsewhere [14,19]. Briefly, the difference in AT between each of the four adjacent electrodes was measured, and the greatest value was plotted as the local maximal phase difference in that area. Local conduction velocity was then calculated as the mean of two conduction vectors in a 2.5×2.5 mm^2^ area. The area covered by the array of electrodes were used to access the regional abnormal conduction parameters (CB, CD and CDCB). As defined in other publications [13,20], CB, CD and CDCB are local conduction velocities as <0.1 m/s, 0.1≤x<0.2 m/s, and <0.2 m/s, respectively.

### Postoperative Atrial Fibrillation

The POAF was defined as new-onset AF within 1 – 4 days of cardiac surgery and prior to hospital discharge.

### Preoperative P-wave Duration

The most recent electrocardiogram (ECG) before cardiac surgery was analyzed for each patient to measure preoperative p-wave duration in Muse 8 (GE Medical Systems Information Technologies, Inc., Milwaukee, WI). The p-wave duration for all 20 patients participating in the study was quantified with a lead II median. Here the term ‘median’ represents the medium of available sinus beats for each patient in the electronic record. Two independent observers evaluated ECGs, and the results were consulted with a cardiac electrophysiologist.

### Statistical Analysis

Demographic comparisons between patients who did and did not develop POAF were performed using the student t-test. The data are expressed as mean±SD. The student t-test was used for normally distributed parametric variables. For paired data set evaluation, a non-parametric test (Wilcoxon matched-pairs signed rank) was used where the distribution was not normal. For unpaired data sets with normal distribution Kolmogorov-Smirnov and Shapiro-Wilk tests were used. The Mann-Whitney U test was used for non-normally distributed variables. PRISM 8.0 (GraphPad Software, Inc., San Diego, CA) was used for all data analysis. Statistical significance was set at a p<0.05 by 2-tailed test.

## RESULTS

### Study Population

The baseline characteristics, preoperative medications, and preoperative and intraoperative clinical parameters of cardiac surgery patients are summarized in **Table 1**. Six patients (30%) developed POAF with an average age of 71±6 years which is trending towards being significant in comparison with those who remained in sinus after cardiac surgery, 57±16 years, p=0.054.

### Epicardial High-Resolution Activation Time Mapping Data

Two adjacent electrodes located at the center of the array was used for premature S1S2 electrical pacing (**Fig. S1**). Examples of AT maps from patients (include some details for SR patients with CD, CB, CDCB) that remained in sinus after cardiac surgery (**Fig. 2a**) and in patients who developed POAF (**Fig. 2b**) are shown. The upper panel of **Fig. 2a** and **2b** shows the AT maps for the last S1 beat of consecutive 8 beats with each CL of 600 ms, and the lower panel of **Fig. 2a** and **Fig. 2b** depicts the AT maps recorded for a premature atrial S2 beat with the shortest CL captured.

**Fig. 2.**
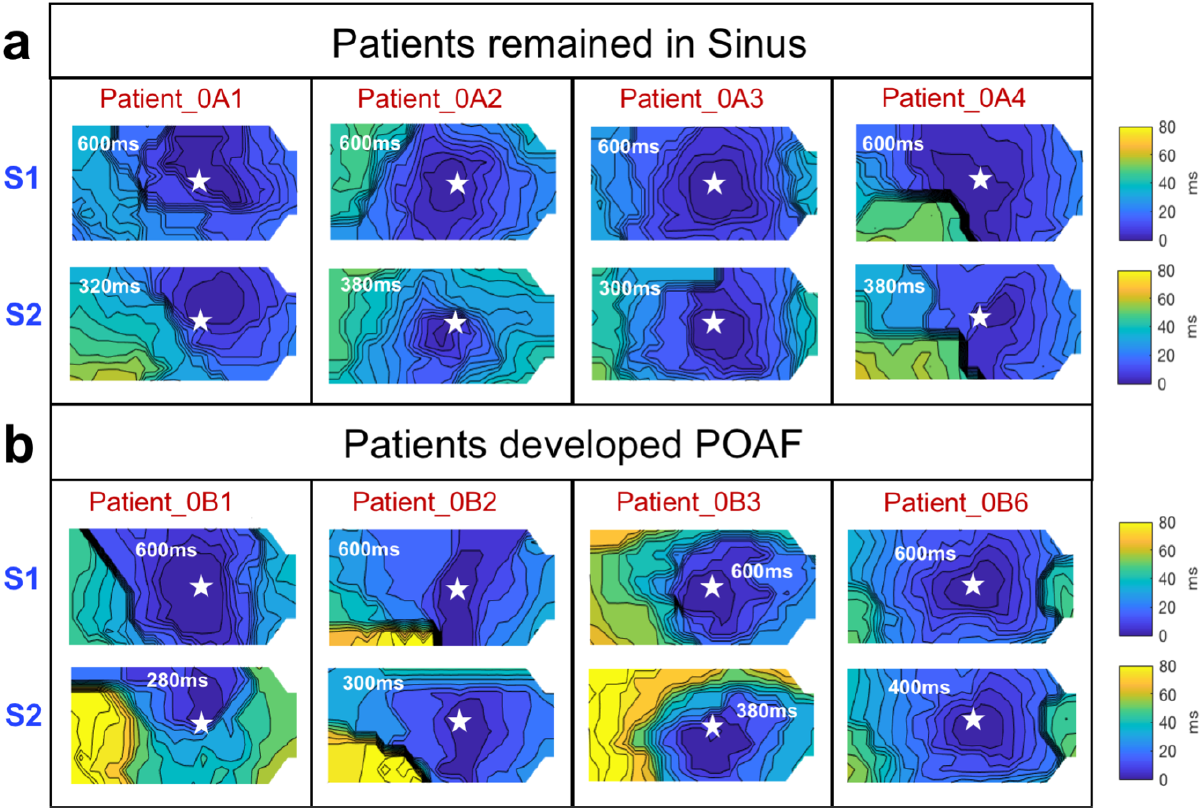
High-density intraoperative LA electrical AT maps during S1S2 stimulation in cardiac surgery patients. **(a)** Data set of five patients who remained in sinus after cardiac surgery. **(b)** Data set of five patients who developed POAF condition within 1 – 4 days of cardiac surgery. **(Top)** AT maps for the last S1 atrial captured beat of 8 consecutive beats with each CL of 600 ms. **(Bottom)** AT maps recorded for a premature atrial beat S2 with the shortest CL. Star (★) represents the site of pacing electrodes.

### Conduction Disorder Quantification During S1S2 Pacing

Examples of conduction velocity maps for a premature atrial S2 beat with the shortest CL captured at the LA posterior wall of three patients that remained in sinus following cardiac surgery and another three patients that developed POAF condition within 1 – 4 days of cardiac surgery are shown in **Fig. 3a** and **Fig. 3b**, respectively. The patients who developed POAF exhibit a greater area of CB, CD and CDCB in comparison to those who remained in sinus. Further, as shown in Fig. 3b, patients that developed POAF have irregular and spatially distributed small patches for CD and CDCB, in comparison to those who remained in sinus (**Fig. 3a**).

**Fig. 3.**
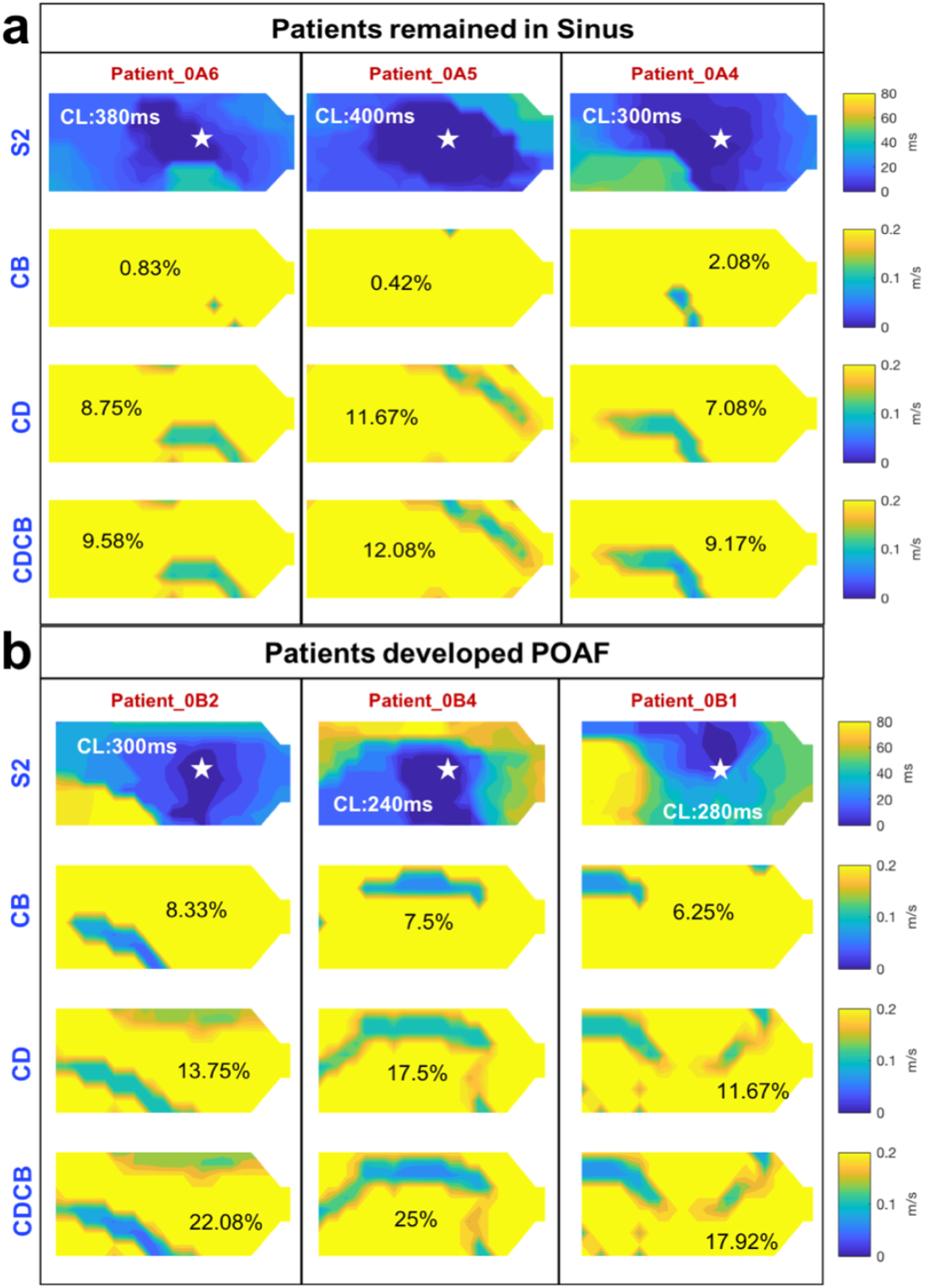
Conduction electrograms show areas relevant to CB, CD and CDCB for a premature atrial beat S2 in three patients that remained in sinus **(a)** and in three patients that developed POAF **(b)**. CB, CD and CDCB refer to spatial distribution of local CV (m/s) as <0.1, 0.1≤x<0.2 and <0.2, respectively. Incidence of conduction disorders is lower in patients that remained in SR in comparison to those who developed POAF. CB, CD and CDCB were assessed for AT maps recorded for premature atrial stimulation S2. Star (★) represents the site of pacing electrodes.

The conduction velocity maps for both premature S1 and S2 beats were quantified for conduction disorders as CB: <0.1 m/s, CD: 0.1≤x<0.2 m/s and CDCB: <0.2 m/s to examine if the premature atrial S2 beats accentuate conduction abnormalities in cardiac surgery patients and reveal a pre-surgical atrial substrate that may indicate greater risk for POAF. **Fig. 4** shows the quantified conduction disorder data as CB (**Fig. 4a**), CD **(Fig. 4b)** and CDCB (**Fig. 4c**) for both S1 and S2 beats in group of patients that developed POAF in comparison to those who remained in sinus following cardiac surgery. For S1 atrial captured beats, the differences in areas captured as conduction abnormalities based-on the mapped region between the groups of patients (POAF versus sinus) are not significant for CB (1.32±2.16 versus 0.56±1.18 mm^2^, p=0.324), CD (6.39±5.93 versus 3.33±3.36 mm^2^, p=0.281) and CDCB (7.71±6.92 versus 3.89±4.39 mm^2^, p=0.258). Unlike S1 atrial beat, the premature atrial S2 beat with the shortest CL captured shows a significantly higher area for CD (13.19±6.59 versus 6.06±4.22 mm^2^, p=0.028) and CDCB (17.36±8.75 versus 7.41±6.39 mm^2^, p=0.034), and shows a trend towards significance for CB (4.17±3.66 versus 1.34±2.86 mm^2^, p=0.063) in patients that developed POAF in comparison to those who remained in sinus.

**Fig. 4.**
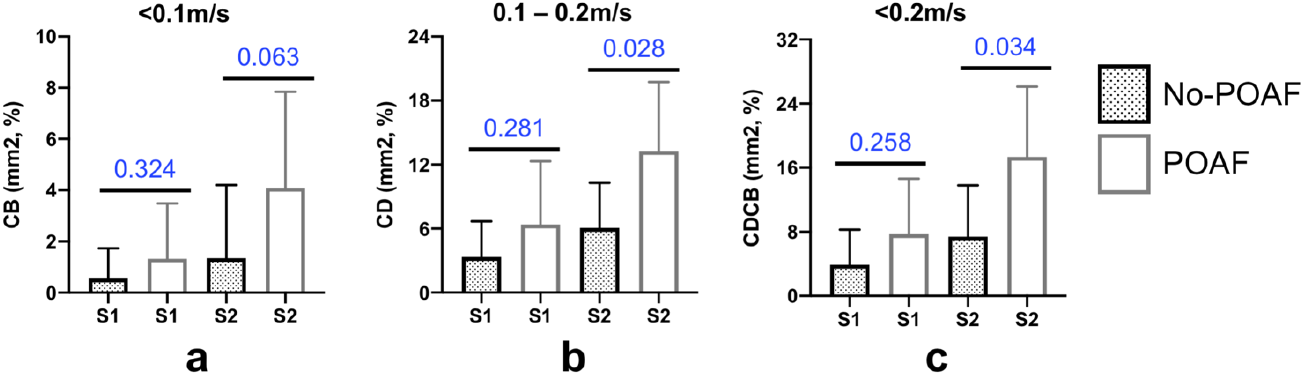
Quantified conduction disorders: **(a)** CB, **(b)** CD and **(c)** CDCB recorded for premature atrial S2 beat with the shortest CL captured and the last S1 atrial captured beat of 8 consecutive beats with each CL of 600 ms. The premature atrial beat S2 captures a larger area of electrodes for CB (4.17±3.66 versus 1.34±2.86, p=0.063), and a significantly much larger area for CD (13.19±6.59 versus 6.06±4.22, p=0.028) and CDCB (17.36±8.75 versus 7.41±6.39, p=0.034) in patients that developed POAF compared to patients that remained in sinus. ‘%’ refers to total area in mm^2^ of electrodes in percentage revealing CB (<0.1 m/s), CD (0.1≤x<0.2 m/s) and CDCB (<0.2 m/s).

### Conduction Disorder Quantification in Sinus

**Fig. 5** shows the recorded AT maps and the quantified conduction abnormalities for normal sinus beats in patients that did not develop POAF (**Fig. 5a**), and in patients that developed POAF (**Fig. 5b**). **Fig. 5c** shows the quantified data representing CB (**left**), CD (**center**) and CDCB (**right**) in each group. The differences in area covered by conduction disorders between the two groups (sinus versus POAF) for CB (0.05±0.14 versus 0.07±0.17 mm^2^, p=0.794), CD (0.47±1.12 versus 0.98±1.01 mm^2^, p=0.189) and CDCB (0.51±1.26 versus1.05±1.08 mm^2^, p=0.184) are not significant.

**Fig. 5.**
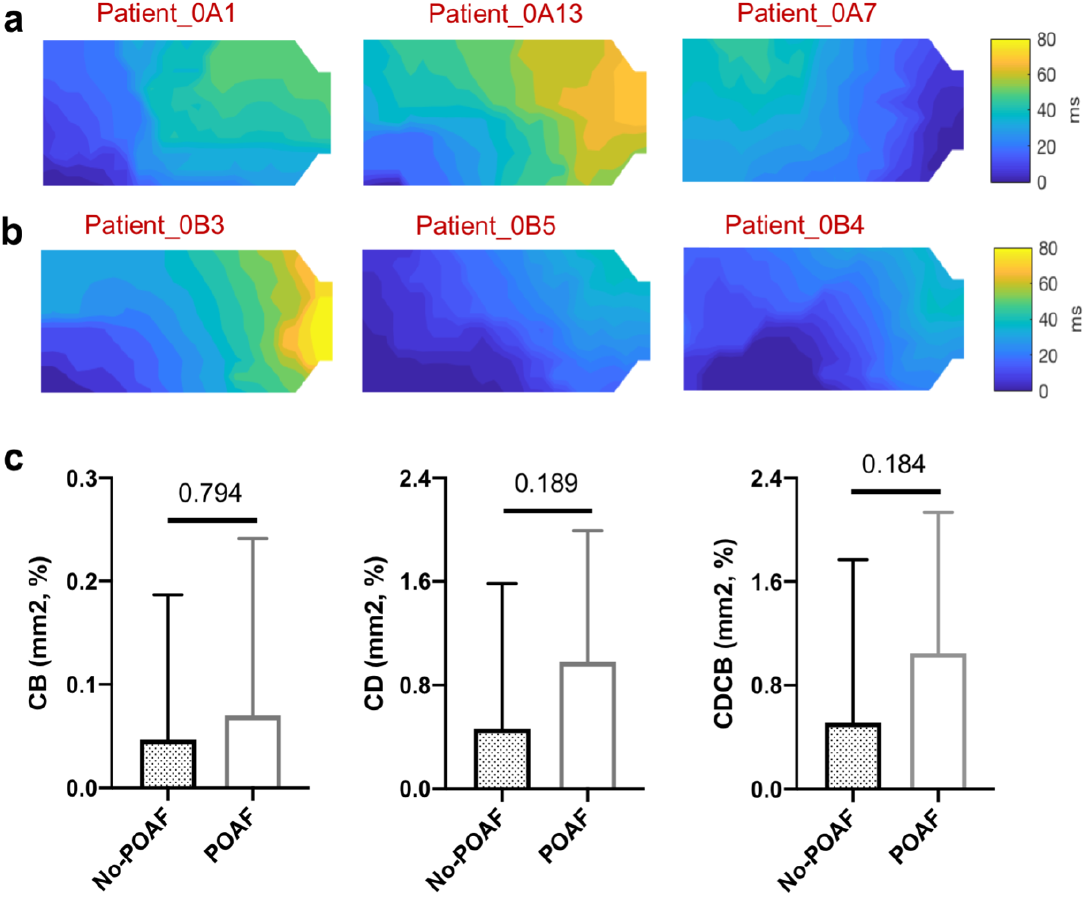
Intraoperative electrical AT data mapped for sinus rhythm in cardiac surgery patients and conduction disorders. **(a)** Example patients that remained in sinus after cardiac surgery. **(b)** Example patients that developed POAF. **(c)** Quantified conduction data for a sinus beats recorded before S1S2 pacing: CB **(left)**, CD **(center)** and CDCB **(right)**.

### Preoperative P-wave Duration

Based on our small cohort patient population (n=20), there is no difference of p-wave duration (109±11 versus 107±11 ms, p=0.799) in patients that developed POAF in comparison with those who remained in sinus.

## DISCUSSION

### Key Findings

This prospective proof-of-concept study of 20 patients recorded and quantified the areas for CB, CD and CDCB at the posterior LA epicardial surface during SR and S1S2 pacing in cardiac surgery patients with no previous history of AF. The current findings demonstrated that unlike SR and S1 beats, premature atrial S2 beats with the shortest CL captured show a significantly higher areas for CD and CDCB, and a trend towards significance for CB in patients that developed POAF in comparison to those who remained in sinus following cardiac surgery.

### Association of Conduction Disorders with POAF Development

Spach et al [16] observed that a premature stimuli resulted in increased dispersion in refractoriness and provoked arrhythmogenic conduction in isolated human atrial bundles. Recently, premature atrial contractions and their role in provoking conduction disorders across atria have been examined in cardiac surgery patients with and without history of AF [17,21]. In these studies, they emphasized that in comparison to sinus recordings, atrial extra-systoles could play a crucial role in quantifying slow and blocked conduction areas observed on the atrial epicardium and endocardium. Similarly, as evident in our results (**Fig. 4**), the premature atrial S2 beats evoked larger areas of CB (**Fig. 4a**), CD (**Fig. 4b**) and CDCB (**Fig. 4c**) in patients that developed POAF in comparison to those that remained in sinus after cardiac surgery.

Lanters et al. [11] demonstrated intraoperative mapping of right atrium (RA), Bachmann’s Bundle and LA in sinus, and reported conduction abnormalities as CB and CD in 209 cardiac patients without a history of AF. Their conduction data for sinus beats do not show any significant correlation between CD and POAF (p=0.650), and CB and POAF (p=0.813). A similar trend was observed at the LA posterior wall in our prospective study where the measured conduction abnormalities recorded during sinus have no association with POAF (**Fig. 5c)**. Similarly, Corina et al. [22] demonstrated conduction maps across LA, RA and Bachmann bundle sites during sinus and unveil obesity-related conduction disorders in obese patients that developed POAF.

The conduction disorder data were previously used to find the association of epicardial breakthrough waves present within muscular connections between endo-epicardial layer with POAF during sinus in 350 cardiac patients [23]. Their results showed that the presence of epicardial breakthrough waves do not predispose for the development of POAF (p=0.732). In their subsequent work [15], they reported intra-operative high-density electrical mapping of the PV area during sinus in 268 cardiac patients with and without prior history of AF. In a sub-set of 70 patients (with no prior history of AF), their conduction data exhibited a trend towards a higher number of continuous CDCB lines (p=0.030) and long CB lines (p=0.073), and these results are not in agreement with our conduction data quantified for sinus beats (**Fig. 5c**). One possible reason for these differences is that the definition of CB and CD is in terms of conduction velocity. For instance, they referred to CD in the range 0.17 – 0.29 m/s and CB as <0.17 m/s. In our study, based-on previously published definition for CB and CD,[13,20] we evaluated CB and CD for <0.1m/s and 0.1≤x<0.2 m/s, respectively.

Further, some discrepancies among studies could be explained by different positions of the electrode array and the contact area covered by the array of electrodes. For instance, studies reported by Lanters et al. [11], Mouws et al. [15] and Roberts-Thomson et al. [13,14] have differences in their array design, interelectrode spacing, and with some variations in positioning the array on the LA wall while recording epicardial atrial AT data in sinus.

To examine the differences between premature atrial S1 beat and S2 beat within the same group, conduction disorders were quantified as presented in **Table S1**. A significant amount of area for CD and CDCB were observed for S2 beats in comparison with S1 beats in both groups (POAF and sinus). This further strength the application of using an intraoperative premature atrial S2 beats in cardiac surgery patients revealing the pre-surgical atrial substrate and its relevant conduction abnormalities.

### Preoperative P-wave Duration

Previously reported prospective studies and meta-analysis showed that preoperative p-wave duration can be used as an independent parameter to predict POAF in cardiac surgery patients. For instance, Wu et al. [24] reported a study of 299 patients with no prior history of AF, and the p-wave duration cut-off value to predict POAF in these patients was 105 ms. In our study, we reported the mean value of 109±11 ms in patients who developed POAF, and the p-wave duration difference is minimal in comparison to those who remained in sinus post-surgery. Interestingly, in our study there was a trend toward a smaller LAd in patients that developed POAF (**Table 1**).

### Clinical Perspective

Unlike sinus beats, premature atrial stimulated beats have shown significant benefits in assessing pre-surgical abnormal conduction areas present at LA wall and their association with POAF. This opens a new horizon in advancing clinical electrophysiology by evaluating the impact of premature atrial stimulus beats at shortest CL on both intra-and post-operative management.

### Study Limitations

The results presented in this proof-of-concept prospective study supports the hypothesis that a premature atrial S2 beat with the shortest CL captured is useful to quantify conduction disorders (CB, CD and CDCB) which are related to POAF initiation. A major limitation is the sample size (n=20) of this proof-of-concept study. Larger, multicenter studies are warranted to see if these findings are reproduceable while accounting for confounding factors such as surgery type (CABG, mitral and/or aortic valve procedures) and age of patients. Though we cannot rule out the significance of premature atrial beat S2 in quantifying conduction disorders at posterior LA wall, the effects of premature atrial stimulation during S1S2 pacing at other sites of atria (RA and Bachmann Bundle) remain unknown and further research is warranted.

## CONCLUSIONS

The results presented in this study supports the hypothesis that a premature atrial S2 beat with the shortest CL captured is useful to quantify conduction disorders (CB, CD and CDCB) which are related to POAF initiation. Conduction abnormalities in patients who remained in sinus and who developed POAF following cardiac surgery were not apparent during sinus and S1 pacing beats. This proof-of-concept study creates a reference dataset for future multisite atrial mapping studies at LA, RA and Bachmann bundle during S1S2 pacing in a large cohort of cardiac surgery patients with no prior history of AF.

## Data Availability

The datasets generated during and/or analyzed during the current study are available from the corresponding author on reasonable request.

## Acknowledgments

D.J.D. conceived the clinical research study. M.S.K. and D.J.D. collected intraoperative atrial activation time data along with clinicians (C.S., J.P.G. and V.S.). M.S.K. performed postoperative atrial activation time data analysis and generated conduction velocity maps. R.R. supervised electrophysiology parameters, and M.S.K. and M.L. measured preoperative p-wave duration as two observers. C.S., J.P.G. and V.S. performed cardiac surgeries. M.S.K. wrote the manuscript and D.J.D. edited the manuscript. All authors approved the final copy.

## Human subjects/informed consent statement

The study protocol was approved by the Institutional Review Board of the University of Utah – Health (IRB_00101187). All experiments were performed in accordance with relevant guidelines and regulations as approved, and the written informed consent was obtained from all 22 patients.

## Additional Information

Supplementary information is available for this paper in the supporting document attached to this file.

---

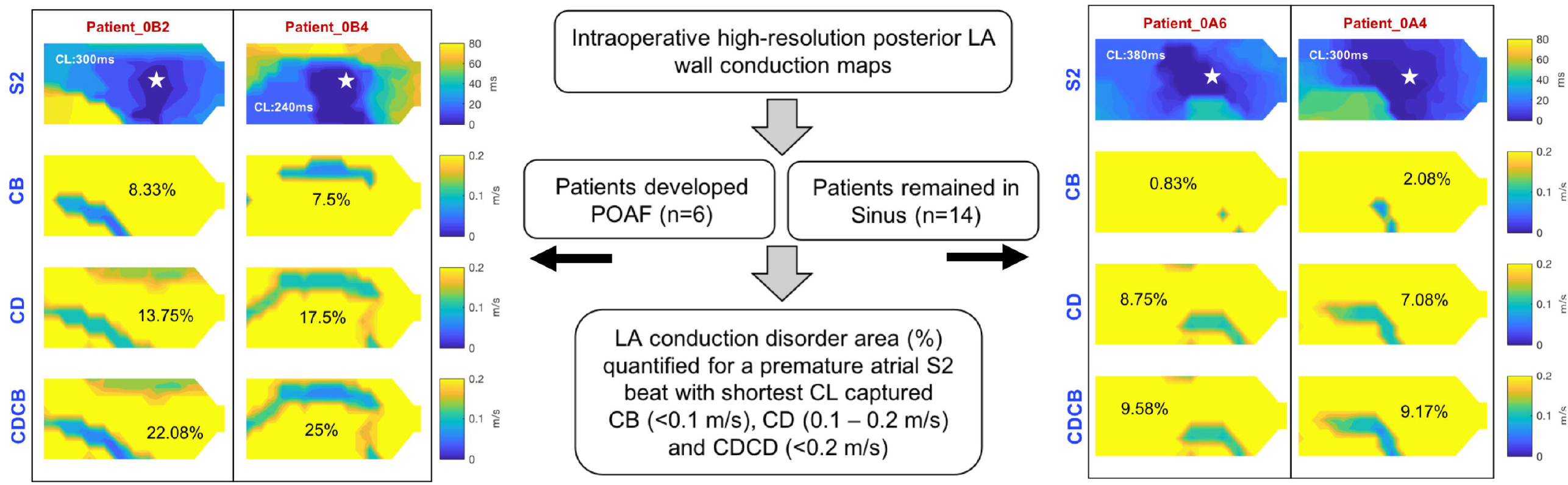

Premature atrial S2 beat with the shortest CL captured reveals a larger area for CB, CD and CDCB in patients that developed POAF **(left panel)** in comparison to those who remained in sinus following cardiac surgery **(right panel)**. CL: Cycle length, CB: Conduction block, CD: Conduction delay, CDCB: Conduction delay and block.

