## Supplemental Fig. 1 and Table S1 for "Premature Atrial Stimulation Accentuates Conduction Abnormalities in Cardiac Surgery Patients that Develop Postoperative Atrial Fibrillation"

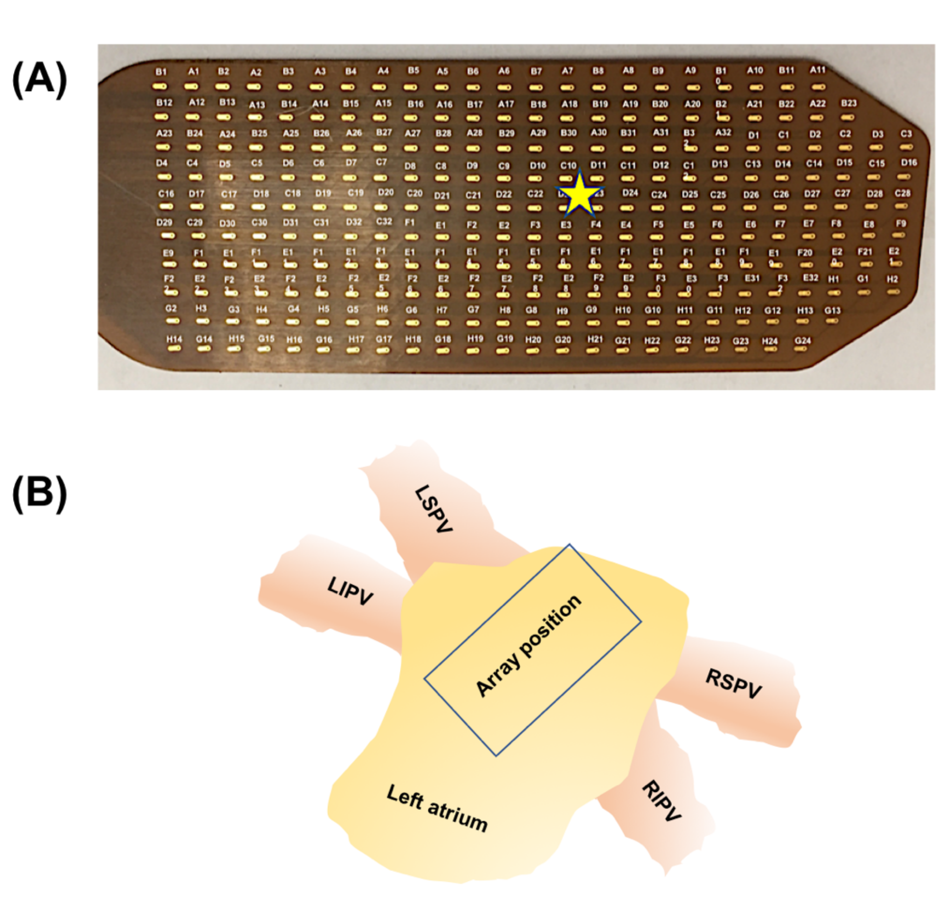


**Fig. S1. (A)** Flexible array of 240-electrodes used in the intraoperative study to collect epicardial atrial activation time data in sinus and S1S2 pacing. **(B)** A schematic representation of an array positioned on posterior LA wall in between PVs (not drawn to scale). Star (★) represents the site of pacing electrodes.

**Table S1.** Comparing effectiveness of a shortest premature captured atrial beat S2 with S1 (600 ms) to quantify CB, CD and CDCDB in patients that remained sinus and developed POAF during 1 – 4 days post-cardiac surgery.

| **Group** | **Conduction disorder type** | **S1 ($mm^2$)** | **S2 ($mm^2$)** | **p value** |
|---|---|---|---|---|
| Sinus | CB | 0.56±1.18 | 1.34±2.86 | 0.126 |
| | CD | 3.33±3.36 | 6.06±4.22 | 0.004 |
| | CDCB | 3.89±4.39 | 7.41±6.39 | 0.004 |
| POAF | CB | 1.32±2.16 | 4.10±3.75 | 0.125 |
| | CD | 6.39±5.93 | 13.26±6.49 | 0.032 |
| | CDCB | 7.71±6.92 | 17.36±8.75 | 0.031 |
